# Beta Burst-Driven Adaptive Deep Brain Stimulation Improves Gait Impairment and Freezing of Gait in Parkinson’s Disease

**DOI:** 10.1101/2024.06.26.24309418

**Authors:** K.B. Wilkins, M.N. Petrucci, E.F. Lambert, J.A. Melbourne, A.S. Gala, P. Akella, L. Parisi, C. Cui, Y.M. Kehnemouyi, S.L. Hoffman, S. Aditham, C. Diep, H.J. Dorris, J.E. Parker, J.A. Herron, H.M Bronte-Stewart

**Author notes:** Corresponding Author: Helen Bronte-Stewart, Department of Neurology and Neurological Sciences, Center for Academic Medicine, Rm 254A, 453 Quarry Rd, Stanford University School of Medicine, Stanford, CA 94305. Co-First Authors (K.B. Wilkins, M.N. Petrucci).

## Abstract

**Background:** Freezing of gait (FOG) is a debilitating symptom of Parkinson’s disease (PD) that is often refractory to medication. Pathological prolonged beta bursts within the subthalamic nucleus (STN) are associated with both worse impairment and freezing behavior in PD, which are improved with deep brain stimulation (DBS). The goal of the current study was to investigate the feasibility, safety, and tolerability of beta burst-driven adaptive DBS (aDBS) for FOG in PD.

**Methods:** Seven individuals with PD were implanted with the investigational Summit™ RC+S DBS system (Medtronic, PLC) with leads placed bilaterally in the STN. A PC-in-the-loop architecture was used to adjust stimulation amplitude in real-time based on the observed beta burst durations in the STN. Participants performed either a harnessed stepping-in-place task or a free walking turning and barrier course, as well as clinical motor assessments and instrumented measures of bradykinesia, OFF stimulation, on aDBS, continuous DBS (cDBS), or random intermittent DBS (iDBS).

**Results:** Beta burst driven aDBS was successfully implemented and deemed safe and tolerable in all seven participants. Gait metrics such as overall percent time freezing and mean peak shank angular velocity improved from OFF to aDBS and showed similar efficacy as cDBS. Similar improvements were also seen for overall clinical motor impairment, including tremor, as well as quantitative metrics of bradykinesia.

**Conclusion:** Beta burst driven adaptive DBS was feasible, safe, and tolerable in individuals with PD with gait impairment and FOG.

## Introduction

Gait impairment and freezing of gait (FOG) can lead to falls, injury, and loss of independence for individuals with Parkinson’s disease (PD)^1,2^. Despite the efficacy of dopaminergic medication for symptoms such as bradykinesia and rigidity, gait impairment and FOG may be refractory to medication^3,4^. Traditional continuous deep brain stimulation (cDBS) improves gait impairment and FOG to a certain degree in individuals with PD who are responsive to medication^5^, but many patients still experience these symptoms on DBS despite effective treatment of other symptoms, and often gait continues to decline over time^6–9^. Therefore, there is a growing need for novel therapies that may better target gait impairment and FOG in PD.

Subcortical local field potential (LFP) recordings from the subthalamic nucleus (STN) revealed a potential pathological neural feature of FOG in PD^5^. Freezers showed prolonged beta band (13-30 Hz) burst durations compared to non-freezers in the STN, which further increased during episodes of FOG. DBS acted to shorten these pathologically long beta bursts, and improved FOG. Considering beta burst duration both relates to the observed impairment of FOG and is also modulated by DBS intensity alongside improvements in behavior, it offers the potential to serve as a relevant neural input for adaptive DBS (aDBS) in which stimulation amplitude is adjusted in real-time based on a given input. To date, the majority of aDBS studies have been implemented either at rest, during seated tasks, or in patients who did not exhibit FOG, and therefore it is unknown how aDBS may affect gait impairment and FOG. An initial case study using the first-generation sensing neurostimulator (Activa® PC+S, Medtronic, PLC) evaluating beta power-driven aDBS found improved gait and reduced FOG compared to OFF DBS and traditional cDBS in one participant based on quantitative kinetics^10^. However, this device was unable to use beta burst durations as a neural input due to technical limitations and was only in one participant.

Advances in DBS technology now enable the possibility of using beta burst durations as a neural input for aDBS in the chronic setting using the investigative Summit™ RC+S neurostimulator (Medtronic, PLC). The goal of the current study was to investigate the feasibility, safety, and tolerability of beta burst-driven aDBS for gait impairment and FOG in PD. We evaluated quantitative assessments of gait impairment and FOG, as well as other PD motor symptoms in a blinded, randomized study.

## Materials and Methods

### Human Subjects

Seven participants (5 male, 2 female) with clinically established PD underwent bilateral implantation of DBS leads (model 3389, Medtronic, PLC) in the sensorimotor region of the STN using a standard functional frameless stereotactic technique and multi-pass microelectrode recording^11^. The two leads were connected to the implanted investigative neurostimulator Summit™ RC+S (Medtronic, PLC). All implantable pulse generators (IPGs) were implanted in the right chest. Inclusion criteria included being at least 18 years of age, meeting criteria for STN DBS^11^, presence of complications of medication such as wearing off signs, fluctuating responses, dyskinesias, medication refractory tremor, and/or impairment in the quality of life on or off medication, and a score ≥ 1 on the Freezing of gait questionnaire (FOG-Q) and/or gait sub-score (Item 3.10) of Movement Disorders Society Unified Parkinson’s Disease Rating Scale Part III (MDS-UPDRS III). Exclusion criteria included being over the age of 80, dementia, untreated psychiatric disease, Hoehn and Yahr stage 5 (non-ambulatory), major surgical morbidities such as severe hypertension, coagulopathy, or conditions that might increase the risk of hemorrhage or other surgical complications, presence of a cardiac pacemaker, required rTMS, ECT, MRI, or diathermy, pregnancy, cranial metallic implant, or history of seizures or epilepsy. All participants gave written consent to participate in the study, which was approved by the Food and Drug Administration with an Investigational Device Exemption and by the Stanford University Institutional Review Board.

### Experimental Protocol

All experiments occurred after the participant had been clinically optimized on their DBS settings. Experimental testing was done in the off-medication state, which entailed stopping long-acting dopamine agonists at least 48 hours, dopamine agonists and controlled release carbidopa/levodopa at least 24 hours, and short acting medication at least 12 hours before testing. The protocol was split into two separate visits, each consisting of three to six days of testing that were separated by three months. The design of the protocol was a ‘bench to bedside’ model. During the first visit, a harnessed gait task was performed to test the initial safety and tolerability of the neural aDBS settings. If the first visit was successful, then during the second visit, a free walking task was performed. The details of the two visits are as follows:

Both visits proceeded in a similar manner. Participants initially completed OFF DBS testing. For the first visit, the OFF DBS testing consisted of a 100-second stepping-in-place (SIP) task and MDS-UPDRS III by a certified rater. The SIP task is a validated assessment of FOG in PD and involves self-paced alternating stepping on two force plates while harnessed^12^. For the second visit, the OFF DBS testing consisted of the turning and barrier course (TBC), MDS-UPDRS III, and a repetitive wrist flexion-extension (rWFE). The TBC task is a validated free walking task designed to elicit FOG and is made up of various dividers that simulate narrow hallways and doorways^13^. Participants walk in two ellipses in one direction, followed by two figures of eights. They then repeat in the opposite starting direction; the initial direction was randomized. The rWFE task is a validated assessment of bradykinesia^14–17^. During the task, participants flex the forearm so that the elbow is angled at 90° and then flex and extend the hand at the wrist joint as quickly as possible for 30 seconds.

Calibration testing followed the OFF DBS testing for each visit. Randomized stimulation titrations were performed to determine both the therapeutic window for neurostimulation and the corresponding LFP beta band burst duration threshold for neural aDBS. The participant performed either 30 seconds of SIP or one ellipse and one figure of eight in both directions of the TBC at randomized stimulation intensities ranging from 25% to 125% of their clinical stimulation amplitude. Following the stimulation titrations, the participant underwent ramp rate testing to find a tolerable rate for adjusting stimulation during aDBS. The last portion of the calibration testing involved calibration runs of aDBS to test out the determined therapeutic window, ramp rate, and aDBS thresholds. For the SIP visit, this consisted of a 100 second seated rest, 30 second standing rest, and a 100 second SIP run. For the TBC visit, this consisted of a 100 second seated rest, 30 second standing rest, and then one ellipse and one figure of eight in each direction.

Following the calibration testing, participants underwent testing of 3 stimulation conditions: cDBS, aDBS, and random intermittent DBS (iDBS). During cDBS stimulation amplitude is held constant during the trial, whereas during iDBS stimulation amplitude varies randomly using the same ramp rate as aDBS. The order of these conditions was randomized, and the participant was blinded to the condition.

For the SIP visit, there was a 20-minute wash-in period for a given stimulation condition. The participant then completed a 100 second SIP followed by an MDS-UPDRS III conducted by a blinded certified rater. After the MDS-UPDRS III, the participant was asked a customized questionnaire about their experience on that stimulation condition. This was repeated for each of the three stimulation conditions. The three stimulation conditions were either completed in one day or split between two days.

The TBC visit consisted of two timepoints. The stimulation first washed in for 60 minutes. After 60 minutes of a given stimulation condition, the participant completed the TBC task (two ellipses and two figures of eights in both directions), the MDS-UPDRS III by a blinded certified rater, rWFE, and the customized questionnaire. After an additional 60 minutes of a given stimulation condition (i.e., at the 120-minute timepoint), the participant repeated the same tasks and questionnaire. This was repeated for each of the three stimulation conditions. The three stimulation conditions were completed across two to three days.

### Distributed Neural Adaptive DBS System

The current study used the Summit™ RC+S DBS system (Medtronic, PLC). This system consists of a rechargeable implanted neurostimulator (INS) with sensing and closed-loop stimulation capabilities, a bidirectional communicator, and a C# API that allows the creation of custom applications (Figure 1). The API can be used to both configure the implantable neurostimulator (INS) and perform ‘distributed’ closed loop stimulation where neural data is streamed from the INS via Bluetooth to a PC-in-the-loop. The neural data are then analyzed in real-time and commands to change stimulation based on the control policy algorithm are sent back to the INS via the communicator.

**Figure 1.**
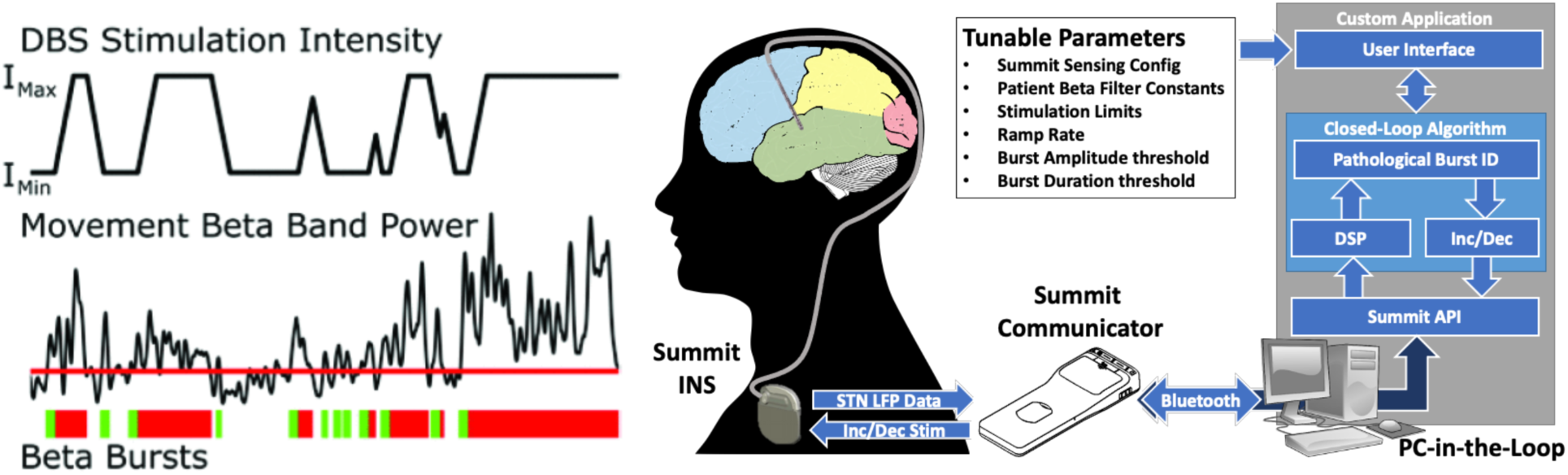
Beta burst duration control policy algorithm and system block diagram. (Left) Burst durations are calculated from sequential crossings of the envelope of beta power over a participant-specific threshold (horizontal red line). Green bars indicate physiological burst durations and the transition to red shows when the burst has become pathological (i.e., exceeded the participant -specific burst duration threshold). DBS intensity increases or decreases at tolerable ramp rates within the therapeutic window in response to the beta burst duration. (Right). PC-in-the-loop architecture utilizes the Summit communicator to provide STN beta burst-based adaptive stimulation. DSP: Digital Signal Processing; Inc/Dec: Increment/Decrement Stimulation. Summit Communicator and INS images are both credited to Medtronic. Figure adapted from Petrucci et al., 2020.

### Neural Adaptive DBS Control Policy Algorithm

A single threshold algorithm based on STN beta burst durations was used, in which stimulation increased when the observed beta burst duration was longer than a participant-specific threshold, or decreased if it was shorter than the threshold^18^. In this algorithm, local field potential (LFP) data were streamed from each STN and added into two buffers. The LFP data were filtered with a 128- order bandpass FIR filter around a participant-specific 6 Hz band within the beta band (13-30 Hz). The filtered data were squared, and then peaks were identified and then linear interpolated to create a beta envelope. A participant-specific threshold for finding the start/end of a given beta burst was determined based on the average trough (minima) power in the linear envelope of the signal in the 45-65 Hz band from the OFF recording^18,19^. The most recent beta burst was identified by finding the period in the buffered data where the envelope was above the participant-specific threshold. The duration of this burst was then compared to the participant-specific beta burst duration threshold. If the duration of the current burst was longer than the participant-specific burst duration threshold, then stimulation was increased, whereas if it was shorter, then stimulation was decreased (Figure 1). This algorithm is based on the concept of identifying ‘pathological’ bursts in which the observed duration is longer than a ‘normal’ physiological burst, which can be determined based on the participant-specific 1/f distribution^19^. Increases and decreases in stimulation were in increment steps or decrement steps of 0.1 mA. This algorithm was implemented bilaterally if both STNs had usable neural signal, with the decision for each STN occurring independently.

### Therapeutic Window

The randomized stimulation intensity titrations during the calibration testing were used to determine the range in which stimulation was allowed to adjust within, i.e., the ‘therapeutic window’ for each participant^20^. The minimum allowed stimulation intensity (i.e., I_min_) was established as the minimum stimulation amplitude that provided an acceptable therapeutic benefit to the participant’s gait compared to the OFF condition. The maximum allowed stimulation intensity (i.e., I_max_) was established as the maximum stimulation amplitude that did not elicit adverse side effects or lead to a deterioration of gait performance. It did not exceed 125% of the participant’s clinical stimulation amplitude. For tremor dominant individuals, the I_min_ was set to a high enough stimulation amplitude in which there was both a therapeutic benefit to gait and observed tremor was largely absent.

### Threshold Determination

The threshold for ‘pathological’ beta burst durations was determined on a participant-specific basis. The average observed beta burst duration during the OFF gait task or at I_min_ was used as the initial threshold for the first calibration run. The threshold was then adjusted based on a combination of observed stimulation modulation, participant feedback, and the observed kinematics during the calibration runs.

### Ramp Rate

Tolerability of ramp rate of stimulation intensity was tested through a custom C# application prior to the calibration run of aDBS^21^. During ramp rate testing, stimulation intensity randomly fluctuated between I_min_ and I_max_ at a given ramp rate for several minutes while the participant was seated at rest. The participant gave feedback of whether they felt any symptoms or sensations such as tingling (e.g., paresthesias). A 0.1 mA/second ramp up and 0.05 mA/second ramp down was used if it was tolerable for the participant. This slow ramp rate was used due to the presence of artifact in the LFP signal after each increment or decrement of stimulation^22^ (See Supplementary Figure 1). The 0.1 mA/second ramp rate allowed sufficient time for the artifact to subside and collection of artifact-free data after a change in stimulation before the next decision was made. If the participant was unable to tolerate the 0.1 mA/sec ramp up, then the ramp was slowed until a tolerable rate was found. The ramp down was always set to half the rate of ramp up to bias stimulation up^23^.

### Parameters for Other DBS Conditions

The calibration runs were also used to establish the parameters used for the cDBS and iDBS conditions. The goal of the cDBS was to match the total electrical energy delivered (TEED) during aDBS. Therefore, the average observed stimulation intensity during the calibration aDBS run was used as the stimulation intensity for the cDBS condition. The goal of the iDBS was to match both the TEED and the general overall pattern of stimulation variation, without linking the stimulation adaption to beta burst durations. Therefore, the observed modulation of stimulation amplitude during the calibration aDBS run was randomly shuffled for the iDBS condition. All three conditions (aDBS, cDBS, and iDBS) used the same active stimulation contacts, frequency (140.1 Hz), and pulse width (60 μs).

### Data Acquisition and Analysis

#### Kinetic and Kinematic Data Acquisition

Ground reactions forces from dual force plates were sampled at 1000 Hz using a Bertec system (Bertec Corporation, Columbus, OH, USA) during the SIP task.

Kinematics were measured during the gait (SIP and TBC) and rWFE tasks using wearable inertial measurement units (IMUs, APDM, Inc., Portland, OR). 11 IMUS sensors were positioned on the body for the gait tasks: 2 on the shanks, 2 on the feet, 2 on the thigh, 1 on the lumbar, 1 on the chest, 2 on the wrists, and 1 on the forehead. 2 IMU sensors were positioned on the dorsum of the hand for the rWFE task. 3D angular velocities from the IMUs’ triaxial gyroscope were sampled at 128 Hz. Video data were collected at 30 frames per second.

#### Local Field Potential Data Acquisition

LFP data were streamed using the investigational Summit™ RC+S neurostimulator. Two channels, one from each STN, were streamed at 500 Hz. A 0.5 Hz onboard high pass filter and two 100 Hz onboard low pass filters were used^24^. Sense-friendly configurations were used, which consisted of flanking or sandwiching the stimulation contacts (0 to 2, 1 to 3, or 0 to 3^20^). If necessary, clinical stimulation contacts were modified to ensure at least one sense-friendly STN configuration. LFPs were defined from the difference between the two sensing contacts, which serves to further reduce stimulation-related artifact through common mode rejection^24,25^. Active recharge was enabled, which reduced stimulation- and ECG-related artifacts. Data were streamed off the device via the CTM in packets averaging 50 ms. Mode 4 was chosen for data streaming, which allowed faster transmission of packets compared to the alternative available Mode 3 on the Summit device. Mode 4 required the CTM to be in closer proximity to the IPG compared to Mode 3. A cloth sleeve held the CTM and was pressed against the IPG using the strap from the chest IMU.

A custom graphical user interface (GUI) application was used for configuring and streaming data based on a C# API provided by Medtronic as part of a research development kit^18^.

#### Synchronization of Local Field Potential and Kinematic Data

Synchronization of neural, kinetic, kinematic, and video recordings was done using internal and external instruments using a data acquisition interface (Power1401) and Spike software (version 2.7, Cambridge Electronic Design, Ltd., Cambridge, England). The APDM IMU system sent a TTL pulse at the start of recording. Bertec force plate data were synchronized using two transient pulses to the force plate simultaneously captured by an accelerometer placed on the force plate connected to the data acquisition interface. Neural data were synchronized using either a 20 Hz 1.5 mA stimulation train for a few seconds in the OFF stimulation condition, or by transiently switching from 140 Hz to 20 Hz and back to 140 Hz in the ON stimulation conditions. Surface electrodes were attached to the skin over the wires of the IPG which connected to the data acquisition interface and allowed detection of the stimulation-induced artifact when switching to 20 Hz.

#### Kinetic and Kinematic Data Analysis Stepping-in-Place (SIP)

Vertical ground reaction forces under each foot were converted to percentage of body weight and then separated into gait cycles. The stride time begins with initial contact of one foot with the ground and ends just prior to contact of the same foot with the ground. Swing time was identified as the duration between when one foot left the force plate and when the same foot contacted the force plate again. Swing times and stride times were used to calculate asymmetry and arrhythmicity. Asymmetry was defined as: asymmetry = 100 × |ln(SSWT/LSWT)|, where SSWT and LSWT correspond to the leg with the shortest and longest mean swing time over the trial. A larger asymmetry value is indicative of more asymmetric gait. Arrhythmicity was defined as the mean stride time coefficient of variation (standard deviation/mean) of both legs. A large stride time coefficient of variation is indicative of less rhythmic gait. A previously validated computerized algorithm was used to identify episodes of FOG from the vertical ground reaction force data^12^.

#### Turning and Barrier Course (TBC)

Angular velocities of the shanks were filtered using a zero-phase 8^th^ order low pass Butterworth filter with a 9 Hz cut-off frequency, and principal component analysis was used to extract the shank angular velocity within the sagittal plane^13^. The sagittal plane angular velocity (SAV) was used to define different aspects of the gait cycle in the TBC task: the beginning of the swing phase was denoted by the positive slope zero crossing; the end of the swing phase was denoted by the subsequent negative zero crossings; and peak shank angular velocities were identified as the first positive peak following the beginning of the swing phases. The time between subsequent zero crossings of the same leg denoted forward swing phase and the time between consecutive peak shank angular velocities was used to calculate stride times. The peaks of shank angular velocity were used to identify strides to avoid difficulty of discerning heel strikes in PD^13^. Peaks were marked as steps only if they exceeded a minimum threshold 10 deg/s for TBC. Swing angular range was calculated by integrating the sagittal angular velocity curve during swing phase. Swing times and stride times were used to calculate asymmetry and arrhythmicity, as defined above for SIP. Episodes of FOG were identified on a stride-by-stride basis using a previously validated logistic regression model^13^. The logistic regression model outputs the freeze probability over time using arrhythmicity and asymmetry over the last six steps, and stride time and swing angular range for the last step as relevant input parameters. A freeze was indicated when the freeze probability from the logistic regression exceeded 0.7.

#### Repetitive Wrist Flexion-Extension (rWFE)

Angular velocity of each hand in the plane of flexion-extension was low-pass filtered using a zero-phase 4^th^ order Butterworth filter with a 4 Hz cut-off frequency. Root mean square velocity (V_rms_) was calculated for 30-s movement window to quantify bradykinesia. Smaller values indicate more severe bradykinesia.

#### Offline Neural Data Analysis

Neural data from the Summit RC+S device are automatically structured into JavaScript object notation (.JSON) format log files by the Summit API. A custom set of MATLAB functions and scripts was used to time align and analyze these files^26^. For the offline neural analysis to determine parameters for aDBS, power spectral density (PSD) estimates were calculated using Welch’s method with a 1-s Hanning window and 50% overlap. PSDs were calculated across the OFF and stimulation titration conditions. Beta bursts were calculated using a 6 Hz band around the peak beta frequency that showed modulation with increasing stimulation intensity using a method described previously^18,19^.

### Total Electrical Energy Delivered

The Total Electrical Energy Delivered (TEED) was calculated using the following equation^27^:

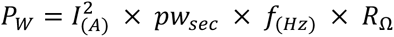

P = power, W = watts, I = current, A = amps, pw = pulse width, f = frequency, R = resistance, and Ω = ohms. In the case of a double monopolar configuration, the TEED was calculated separately for each active contact and then summed. TEED was calculated in one second intervals (TEED (1 s)) and averaged across the trial.

### Lead Reconstruction

Preoperative T_1_ and T_2_ MRI scans and postoperative CT scans were acquired as part of the standard Stanford clinical protocol^11^. Location of DBS leads was determined by the Lead-DBS toolbox^28^. Postoperative CT scans and preoperative T_2_ scans were co-registered to preoperative T_1_ scans, which were then normalized into MNI space using SPM12 (Statistical Parametric Mapping 12; Wellcome Trust Centre for Neuroimaging, UCL, London, UK) and Advanced Normalization Tools^29^. DBS electrode localizations were then corrected for brain-shift in the postoperative CT scan^30^. DBS electrodes were then localized in template space using the PaCER algorithm^31^ and projected onto the DISTAL Atlas to visualize overlap with the STN^32^.

### Statistical Analysis

Statistical analyses were run in R (version 3.6.0, R Foundation for Statistical Computing, University of Auckland, New Zealand). Linear mixed effects models with a fixed effect of stimulation condition and random effect of participant were run for each of the behavioral outcomes. Partial eta squared (*η*_p_^2^) are reported. Estimated marginal means were computed for post hoc pairwise comparisons between stimulation conditions with Tukey’s method to control for multiple comparisons. Significance was set at *p* < 0.05.

## Results

### Demographic Characteristics

Demographic information of the seven individuals with PD is shown in Table 1. The participants (age: 63.7 ± 4.9 years) had a mean pre-op off therapy MDS-UPDRS III of 37.1 ± 9.4 and a disease duration of 14.0 ± 2.5 years. Six of the seven participants were classified as freezers based on the FOG-Q or new FOG-Q (nFOG-Q) and the other participant showed gait impairment as evident by a score of 1 on the MDS-UPDRS III gait item 3.10. Three of the participants were implanted with the Summit RC+S device as part of an IPG replacement, whereas in four participants the Summit RC+S was their initial IPG. Figure 2 visualizes the DBS lead locations within the STN for all participants. Participants performed the SIP visit on average 53.9 ± 32.7 (range: 6-95) months after their Initial Programming and 22.6 ± 9.4 (range: 7-37) months after being implanted with the Summit RC+S. The TBC visit occurred on average 3 ± 1 (range: 2-5) months after the SIP visit.

**Figure 2.**
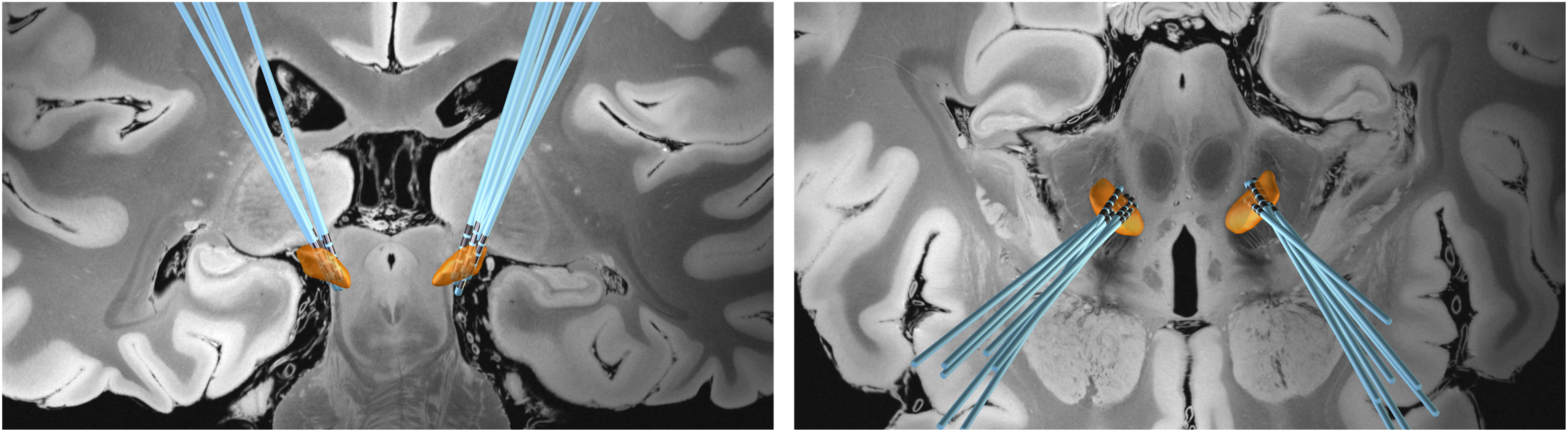
DBS Lead locations within the STN (orange) for all 7 participants. (Left) Coronal and (Right) Axial views.

**Table 1.**
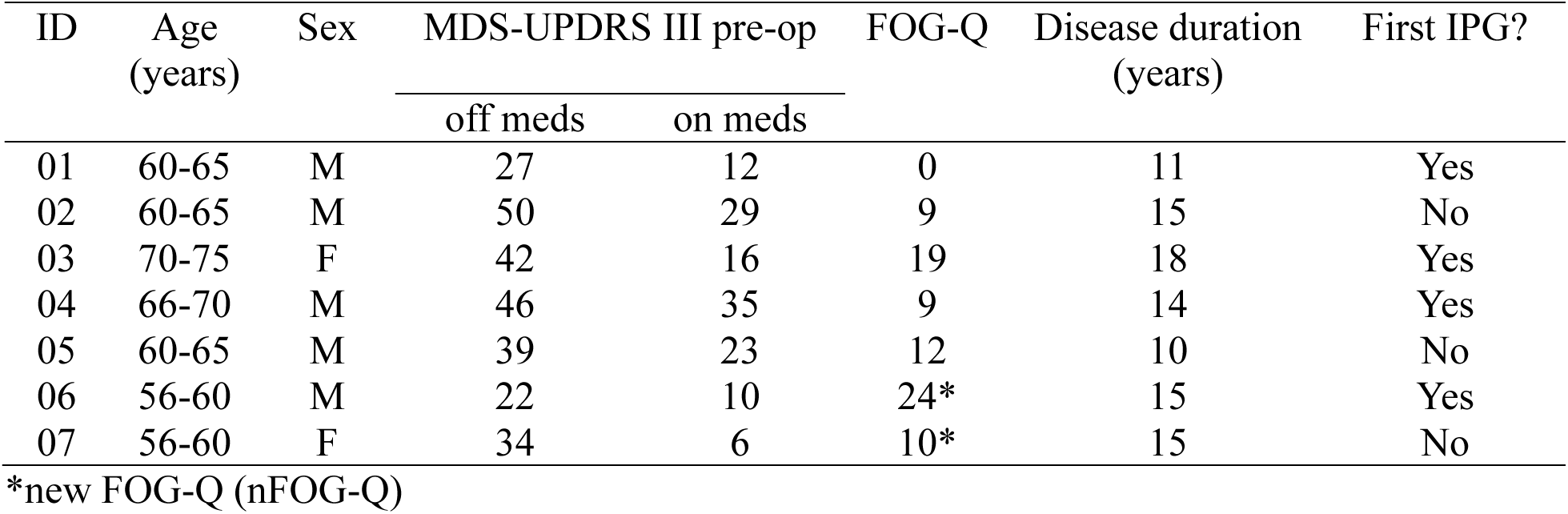
Participant Demographics.

### Calibration of aDBS

Figure 3A shows an example of a participant’s SIP force traces at different stimulation amplitudes as part of the calibration testing. The participant is frozen for the entirety of the trial both at 0% (OFF) stimulation and at 25% of their clinical stimulation amplitude, and for the majority of the trial at 50% clinical stimulation. A noticeable improvement is observed at 75% of their clinical stimulation amplitude, which continues to improve with increasing levels of stimulation up to 110% of their clinical stimulation amplitude. The therapeutic window for this participant was set from 75% to 110% of their clinical stimulation amplitude (Fig. 3B).

**Figure 3.**
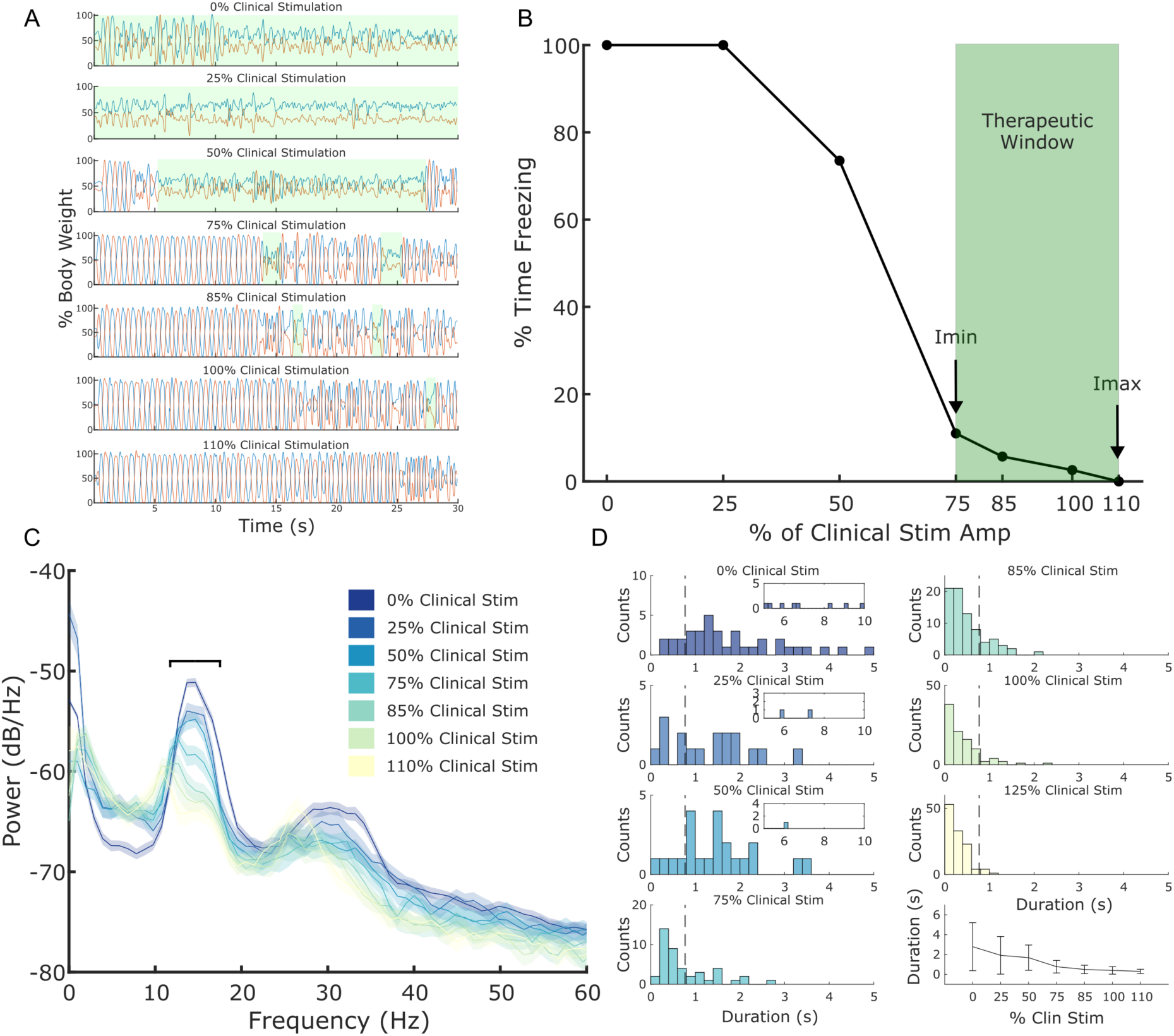
Example of calibration to determine stimulation limits and neural parameters for neural aDBS. (A) Performance during the SIP task at different randomized levels of stimulation amplitude. Stimulation amplitude is represented as % of the participant’s clinical stimulation amplitude. Ground reaction forces from the two force plates are depicted as % body weight. Episodes of FOG highlighted in green. (B) % time freezing during the SIP task at each of the stimulation levels tested with the observed therapeutic window denoted in green. Imin and Imax refer to the minimum and maximum allowed stimulation amplitude during aDBS. (C) Power spectral density (PSD) plots for different levels of stimulation amplitude during the SIP task. Stimulation amplitude is represented as % of the participant’s clinical stimulation amplitude. The chosen 6-Hz band of interest for beta bursts is denoted by the black horizontal line centered around 15 Hz. (D) Histograms of the observed beta burst durations at the different corresponding levels of stimulation amplitude, with the average and standard deviation of the burst duration across stimulation levels in the lower right. The corresponding burst duration threshold is shown by the vertical dashed line.

Figure 3C-D show the matching neural data for one STN from the behavior depicted in Figure 3A-B. A pronounced beta peak is observed around 15 Hz, which attenuates with increasing levels of stimulation intensity. Figure 3D depicts the histograms of the corresponding beta burst durations for the 6-Hz band highlighted in Figure 3C and the corresponding burst duration threshold shown by the vertical dashed line. A wide distribution of beta burst durations is observed in the OFF condition that continues even at 25% and 50% of the participant’s clinical stimulation amplitude, although to a less extreme extent. The burst durations continue to shorten with increasing stimulation amplitude with the majority of the bursts having durations under 500 ms at higher stimulation intensities. Table 2 shows the individual aDBS parameters across the participants at each visit.

**Table 2.** Individual Neural aDBS Controller Parameters.

| ID | Visit | LSTN<br>Contacts | RSTN<br>Contacts | LSTN<br>Range<br>(mA) | RSTN<br>Range<br>(mA) | LSTN<br>Ramp<br>Up /<br>Down<br>(mA/s) | RSTN<br>Ramp<br>Up /<br>Down<br>(mA/s) | LSTN<br>Center<br>Freq<br>(Hz) | RSTN<br>Center<br>Freq<br>(Hz) | LSTN<br>Burst<br>Duration<br>Threshold<br>(ms) | RSTN<br>Burst<br>Duration<br>Threshold<br>(ms) |
| --- | --- | --- | --- | --- | --- | --- | --- | --- | --- | --- | --- |
| 01 | SIP | 1-,2-,C<sup>+</sup> | 9-,C<sup>+</sup> | 2.5-<br>4.9 | 1.8-<br>2.8 | 0.1 /<br>0.05 | 0.1 /<br>0.05 | 23.4 | 17.6 | 225 | 251 |
|  | TBC | 1-,2-,C<sup>+</sup> | 9-,C<sup>+</sup> | 3.6-<br>4.8 | 2.2-<br>2.9 | 0.1 /<br>0.05 | 0.1 /<br>0.05 | 18.5 | 21.5 | 300 | 178 |
| 02 | SIP | 1-,C<sup>+</sup> | 9-,C<sup>+</sup> | 3.7-<br>4.1 | 4.1-<br>4.5 | 0.1 /<br>0.05 | 0.1 /<br>0.05 | 19.5 | - | 400 | - |
|  | TBC | 1-,C<sup>+</sup> | 8-,9-,C<sup>+</sup> | 3.7-4 | 6.4-<br>6.4 | 0.067 /<br>0.033 | - | 17.6 | - | 500 | - |
| 03 | SIP | 1-,2-,C<sup>+</sup> | 9-,C<sup>+</sup> | 2.3-3 | 3.2-<br>3.2 | 0.1 /<br>0.05 | - | 14.6 | - | 771 | - |
|  | TBC | 1-,2-,C<sup>+</sup> | 9-,C<sup>+</sup> | 2.3-<br>3.4 | 3.4-<br>3.4 | 0.1 /<br>0.05 | - | 13.7 | - | 852 | - |
| 04 | SIP | 1-,2-,C<sup>+</sup> | 9-,10-,C<sup>+</sup> | 2.6-<br>3.9 | 5-5 | 0.1 /<br>0.05 | - | 17.5 | - | 270 | - |
|  | TBC | 1-,2-,C<sup>+</sup> | 9-,10-,C<sup>+</sup> | 2.7-<br>4.1 | 5.2-<br>5.2 | 0.1 /<br>0.05 | - | 18.6 | - | 225 | - |
| 05 | SIP | 1-,2-,C<sup>+</sup> | 9-,C<sup>+</sup> | 3.4-<br>4.3 | 3.5-<br>4.5 | 0.1 /<br>0.05 | 0.1 /<br>0.05 | 14.6 | 14.6 | 380 | 350 |
|  | TBC | 1-,2-,C<sup>+</sup> | 8-,9-,C<sup>+</sup> | 4.2-5 | 4.6-<br>5.4 | 0.1 /<br>0.05 | 0.075/<br>0.0375 | 16.5 | - | 390 | - |
| 06 | SIP | 2-,C<sup>+</sup> | 9-,C<sup>+</sup> | 2.2-<br>2.8 | 1.3-<br>1.6 | 0.1 /<br>0.05 | 0.1 /<br>0.05 | 14.7 | 14.6 | 350 | 400 |
|  | TBC | 2-,C<sup>+</sup> | 9-,C<sup>+</sup> | 1.2-<br>2.4 | 0.9-<br>1.7 | 0.1 /<br>0.05 | 0.1 /<br>0.05 | 22.5 | 25.4 | 300 | 254 |
| 07 | SIP | 2-,C<sup>+</sup> | 9-,C<sup>+</sup> | 2.6-<br>3.8 | 4.4-<br>5.9 | 0.075 /<br>0.0375 | 0.075 /<br>0.0375 | 25.3 | 13.7 | 236 | 333 |

\* Dashes indicate instances where an STN was not used for neural recording.

### Stimulation Improves Overall Clinical Motor Impairment

Blinded rated assessments of overall total motor impairment using the MDS-UPDRS III showed a significant effect of stimulation condition (F(3,59.3) = 55.8, p = 2.2e-16, *η*_p_^2^ = 0.74). aDBS (*t* = 11.2, *p* < .0001; 59.9 ± 15.8% improvement), cDBS (*t* = 11.2, *p* < .0001; 58.6 ± 13.8% improvement), and iDBS (*t* = 11.0, *p* < .0001; 57.5 ± 11.7% improvement) all showed significant improvements compared to OFF DBS (Figure 4A). No difference was observed between ON stimulation conditions (aDBS – cDBS: *t* = 0.023, *p* = 1.00; aDBS – iDBS: *t* = 0.19, *p* = 1.00; cDBS – iDBS: *t* = 0.21, *p* = 1.00). One participant was unable to perform the MDS-UPDRS III OFF DBS at the TBC visit due to inability to tolerate being OFF stimulation.

**Figure 4.**
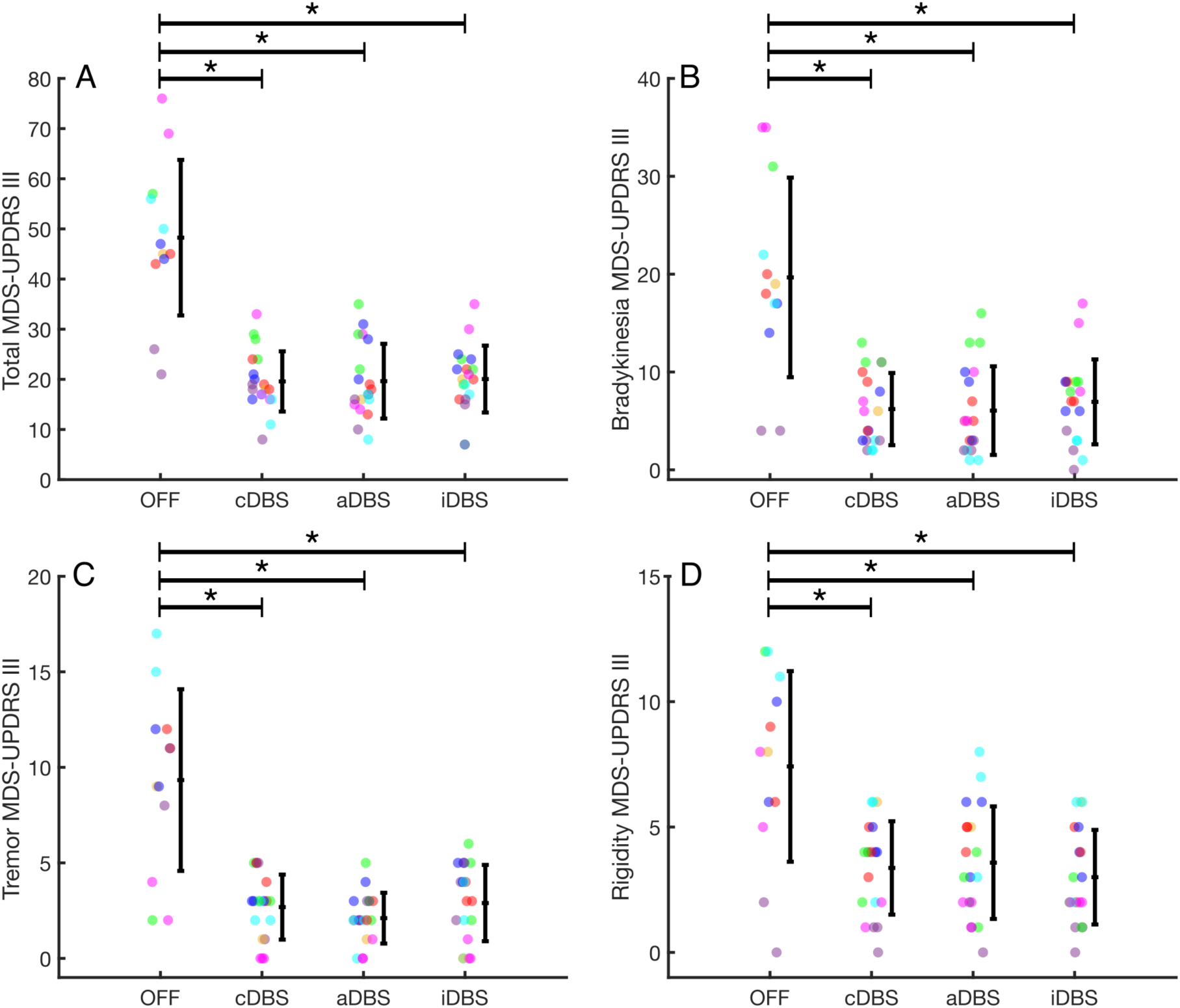
Overall clinical motor impairment and sub-scores across stimulation conditions. (A) Total MDS-UPDRS III, (B) Bradykinesia sub-score, (C) Tremor sub-score, (D) Rigidity sub-score. Individual data are shown, colored by participant ID, along with the average and standard deviation. * indicates p < 0.05.

A significant effect of stimulation condition was seen for each of the sub-scores of the MDS-UPDRS III, including bradykinesia (F(3,59.3) = 40.3, p = 2.60e-14, *η*_p_^2^ = 0.67), tremor (F(3,59.2) = 35.2, p = 3.39e-13, *η*_p_^2^ = 0.64), and rigidity (F(3,59.1) = 20.5, p = 3.28e-9, *η*_p_^2^ = 0.51) (Figure 4B-D). aDBS (bradykinesia: *t* = 9.70, *p* < .0001; tremor: *t* = 9.39, *p* < .0001; rigidity: *t* = 6.25, *p* < .0001), cDBS (bradykinesia: *t* = 9.59, *p* < .0001; tremor: *t* = 8.64, *p* < .0001; rigidity: *t* = 6.60, *p* < .0001), and iDBS (bradykinesia: *t* = 9.08, *p* < .0001; tremor: *t* = 8.37, *p* < .0001; rigidity: *t* = 7.21, *p* < .0001) all showed significant improvements compared to OFF DBS. No difference was observed between any of the ON-stim conditions for any of MDS-UPDRS III sub-scores (see Supplementary Table 1).

### Stimulation Improves Stepping in Harnessed Gait Task

Figure 5 depicts an example neural aDBS trial from the SIP task in one participant. Overall, there was a significant effect of stimulation condition on percent time freezing during the Stepping-In-Place task (F(3,18) = 4.53, *p* = 0.016, *η*_p_^2^ = 0.42). Posthoc pairwise comparisons showed significant reductions in percent time freezing compared to OFF stimulation for aDBS (*t* = 3.22, *p* = 0.022) and cDBS (*t* = 3.01, *p* = 0.035), and a trend for iDBS (*t* = 2.72, *p* = 0.062). No difference was observed between ON stimulation conditions (aDBS – cDBS: *t* = 0.22, *p* = 1.00; aDBS – iDBS: *t* = 0.51, *p* = 0.96; cDBS – iDBS: *t* = 0.29, *p* = 0.99). All five participants who demonstrated freezing in the OFF condition showed improvements in freezing for aDBS and cDBS. In three of those cases, the participant was frozen for the entirety of the trial in the OFF condition and showed substantial improvement with aDBS (25-100% improvement in percent time freezing). The other two participants showed no freezing across any conditions.

**Figure 5.**
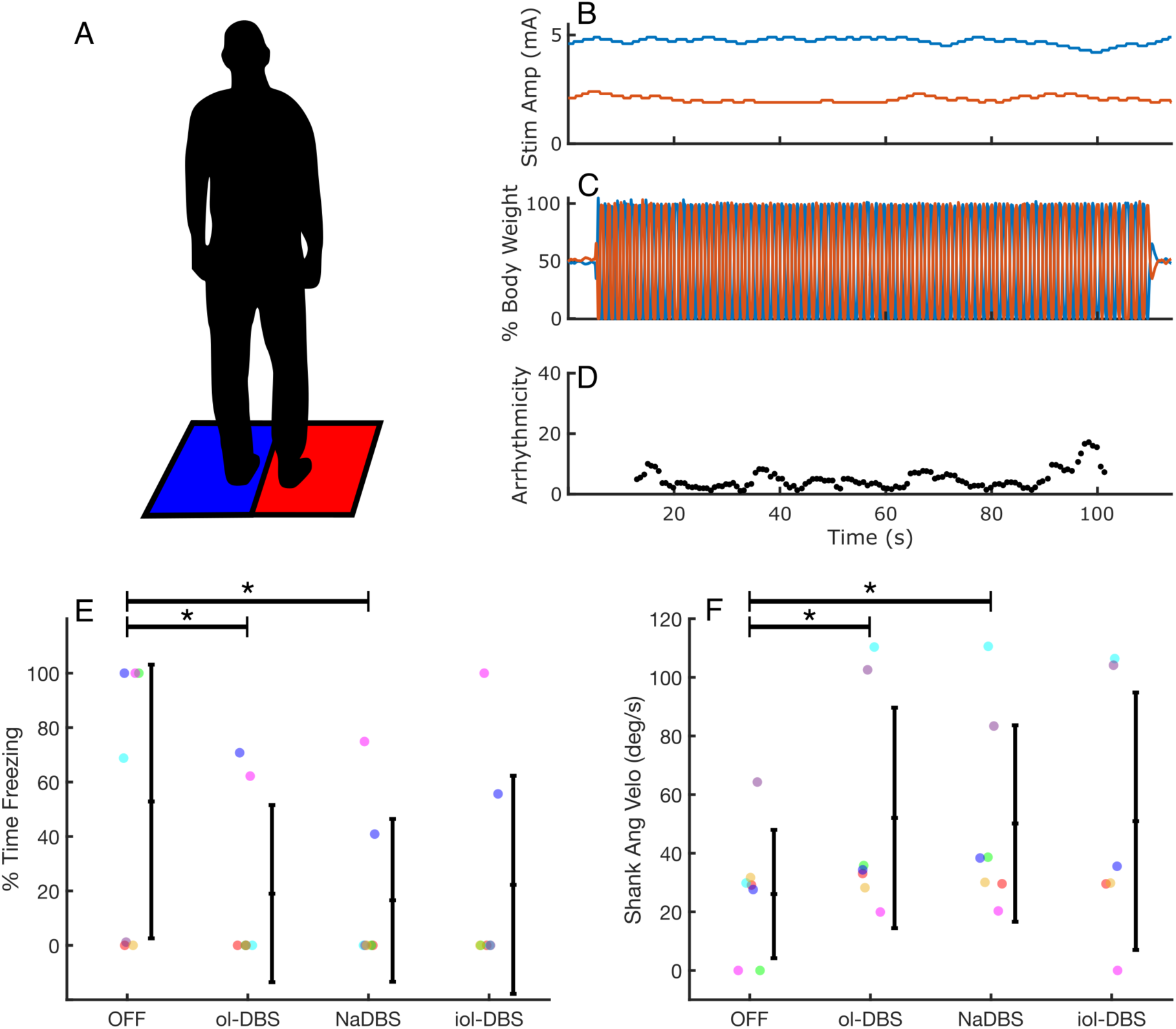
(A) Depiction of the Stepping-In-Place (SIP) task. (B-D) Example of neural aDBS during SIP task for one participant. (B) Stimulation adapting independently for both STNs. The left STN is shown in blue and the right STN in orange. (C) Ground reaction forces from the two force plates depicted as % of body weight. (D) Running arrhythmicity over the trial. (E) % Time Freezing and (F) mean peak shank angular velocity during SIP across stimulation conditions. Individual data are shown, colored by participant ID, along with the average and standard deviation. * indicates p < 0.05.

There was a significant effect of stimulation condition on mean peak shank angular velocity (F(3,17.1) = 4.19, *p* = 0.022, *η*_p_^2^ = 0.42). Posthoc pairwise comparisons showed significant increase in mean peak shank angular velocity compared to OFF stimulation for aDBS (*t* = 2.88, *p* = 0.047) and cDBS (*t* = 3.11, *p* = 0.029), and a trend for iDBS (*t* = 2.52, *p* = 0.093). No difference was observed between ON stimulation conditions (aDBS – cDBS: *t* = 0.23, *p* = 1.00; aDBS – iDBS: *t* = 0.22, *p* = 1.00; cDBS – iDBS: *t* = 0.44, *p* = 0.97). One datapoint for Participant 2 (iDBS) was missing due to an IMU malfunction.

Three participants (Participants 2, 3, and 4) did not have values for the OFF condition for gait arrhythmicity or asymmetry due to being frozen for the entirety of the SIP trial. There was no significant effect of stimulation condition on gait arrhythmicity (F(3,15) = 1.62, *p* = 0.227, *η*_p_^2^ = 0.239). However, it is likely that the lack of data from the three participants who were frozen for the entirety of the SIP trial masked the effect of stimulation condition on gait arrhythmicity, as all three were able to perform the task when ON stimulation. All four participants who were able to step during the OFF condition showed improvements in arrhythmicity for aDBS and cDBS compared to OFF stimulation. There was a significant effect of stimulation condition on asymmetry (F(3,15) = 7.27, *p* = 0.0031, *η*_p_^2^ = 0.59). Posthoc pairwise comparisons showed significant reductions in asymmetry compared to OFF stimulation for aDBS (*t* = 4.59, *p* = 0.0018), cDBS (*t* = 3.57, *p* = 0.013), and iDBS (*t* = 3.55, *p* = 0.014). No difference was observed between ON stimulation conditions (aDBS – cDBS: *t* = 1.25, *p* = 0.61; aDBS – iDBS: *t* = 1.28, *p* = 0.59; cDBS – iDBS: *t* = 0.026, *p* = 1.00). Individual data are included in Supplementary Table 2.

### Stimulation Improves Gait in Free Walking Task

Two participants did not perform the Turning and Barrier Course. Participant 5 did not perform the TBC due to inability to safely perform the task without assistance under clinical stimulation. Although the participant is ambulatory on DBS without assistance, the heightened difficulty of the TBC task due to the narrow hallways and restricted optic flow created by the barriers made it impossible for the participant to complete the task unassisted. They still partook in all other aspects of the visit. Participant 7 did not take part in the TBC visit due to stopping their participation in the trial early for unrelated personal reasons.

Figure 6 depicts an example neural aDBS trial from the TBC task in one participant. Overall, there was a significant effect of stimulation condition on percent time freezing (F(3,27) = 4.73, *p* = 0.0089, *η*_p_^2^ = 0.34). Posthoc pairwise comparisons showed significant reductions in percent time freezing compared to OFF stimulation for aDBS (*t* = 2.95, *p* = 0.031) cDBS (*t* = 3.25, *p* = 0.013), and iDBS (*t* = 3.52, *p* = 0.0080). No difference was observed between ON stimulation conditions (aDBS – cDBS: *t* = 0.47, *p* = 0.97; aDBS – iDBS: *t* = 0.70, *p* = 0.90; cDBS – iDBS: *t* = 0.24, *p* = 1.00). Two of the three participants who demonstrated freezing in the OFF condition showed improvements in freezing for aDBS and cDBS. The two other participants showed no freezing across any conditions.

**Figure 6.**
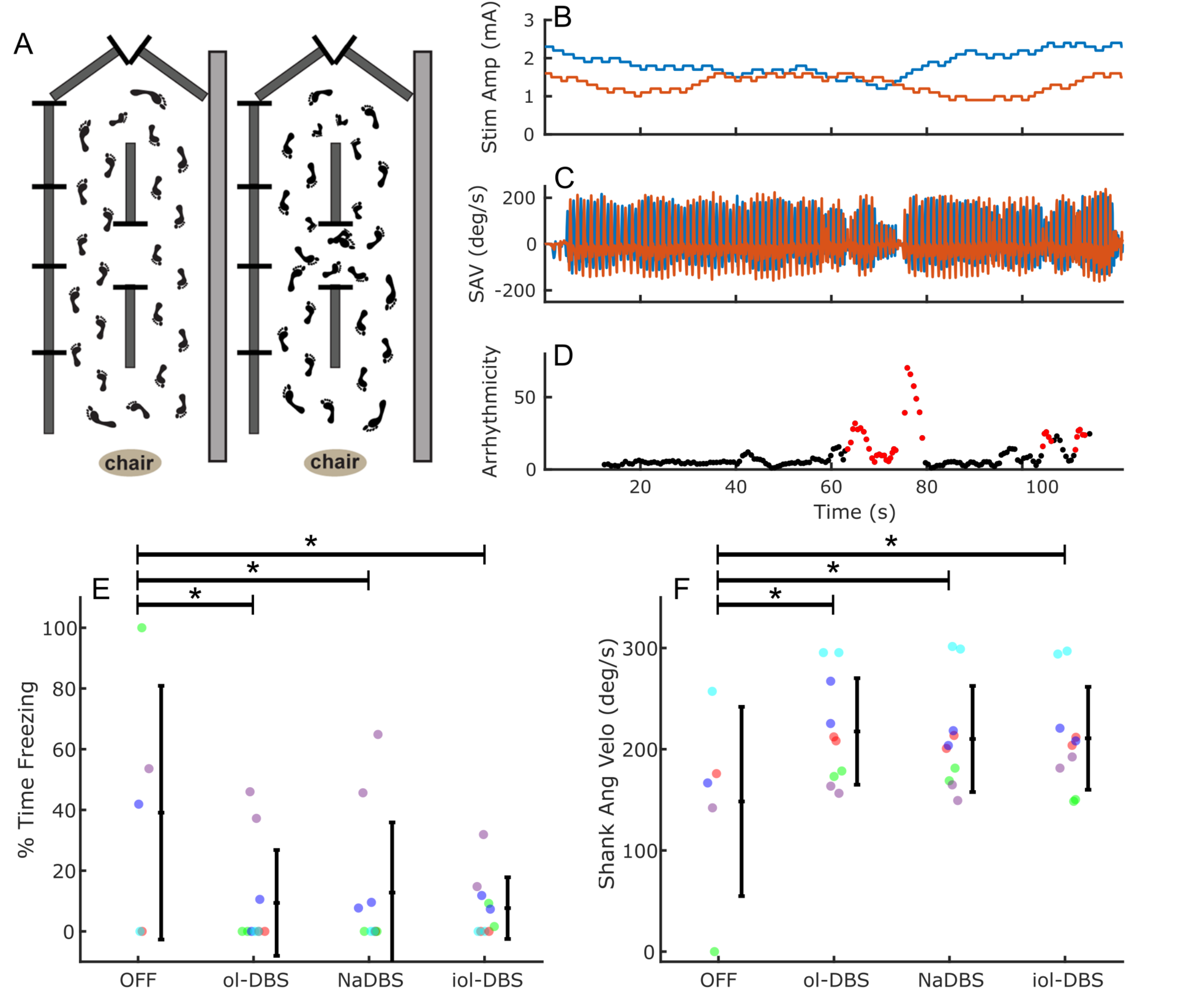
(A) Drawing of a bird’s-eye view of the free walking TBC course showing the ellipses and figure eights that the participant performs. (B-D) Example of neural aDBS during portion of the TBC task for one participant. (B) Stimulation adapting independently for both STNs. The left STN is shown in blue and the right STN in orange. (C) Shank angular velocity for the two legs. (D) Running arrhythmicity over the trial with freezing denoted by the red dots. (E) % Time Freezing and (F) mean peak shank angular velocity during TBC across stimulation conditions. Individual data are shown, colored by participant ID, along with the average and standard deviation. * indicates p < 0.05.

There was a significant effect of stimulation condition on mean peak shank angular velocity (F(3,27) = 9.97, *p* = 0.00013, *η*_p_^2^ = 0.53). Posthoc pairwise comparisons showed significant improvements in mean peak shank angular velocity compared to OFF stimulation for aDBS (*t* = 4.58, *p* = 0.00050), cDBS (*t* = 5.13, *p* = 0.00010), and iDBS (*t* = 4.63, *p* = 0.00050). No difference was observed between ON stimulation conditions (aDBS – cDBS: *t* = 0.68, *p* = 0.91; aDBS – iDBS: *t* = 0.063, *p* = 1.00; cDBS – iDBS: *t* =0.61, *p* = 0.93). All five participants showed improvements in mean peak shank angular velocity for aDBS and cDBS for all timepoints compared to OFF stimulation.

One additional participant (Participant 2) did not have values for the OFF condition for gait arrhythmicity or asymmetry due to being frozen for the entirety of the TBC trial. Overall, there was no significant effect of stimulation condition on gait arrhythmicity (F(3,26) = 0.67, *p* = 0.58, *η*_p_^2^ = 0.072). There was a trend towards an effect of stimulation condition on gait asymmetry (F(3,26.1) = 2,70, *p* = 0.067, *η*_p_^2^ = 0.23). Individual data are included in Supplementary Table 2.

### Stimulation Improves Bradykinesia

Figure 7 depicts an example neural aDBS trial from the WFE task in one participant. Overall, there was a significant effect of stimulation condition on V_rms_ (F(3,70.2) = 24.4, p = 6.42e-11, *η*_p_^2^ = 0.51). Posthoc pairwise comparisons showed significant improvements in V_rms_ compared to OFF stimulation for aDBS (*t* = 7.00, *p* < .0001), cDBS (*t* = 8.24, *p* < .0001), and iDBS (*t* = 6.36, *p* < .0001). No difference was observed between ON stimulation conditions (aDBS – cDBS: *t* = 1.20, *p* = 0.63; aDBS – iDBS: *t* = 1.05, *p* = 0.72; cDBS – iDBS: *t* = 2.36, *p* < 0.094). The second 120-minute timepoint for Participant 3 for the aDBS condition was missing for both hands due to an IMU malfunction. Participant 6 did not perform the rWFE with their right hand at several timepoints (120-minute timepoint for cDBS and both timepoints for aDBS) due to chronic pain in their right shoulder. This pain was unrelated to the stimulation conditions. Five out of the six participants showed improvements on aDBS and cDBS compared to OFF stimulation.

**Figure 7.**
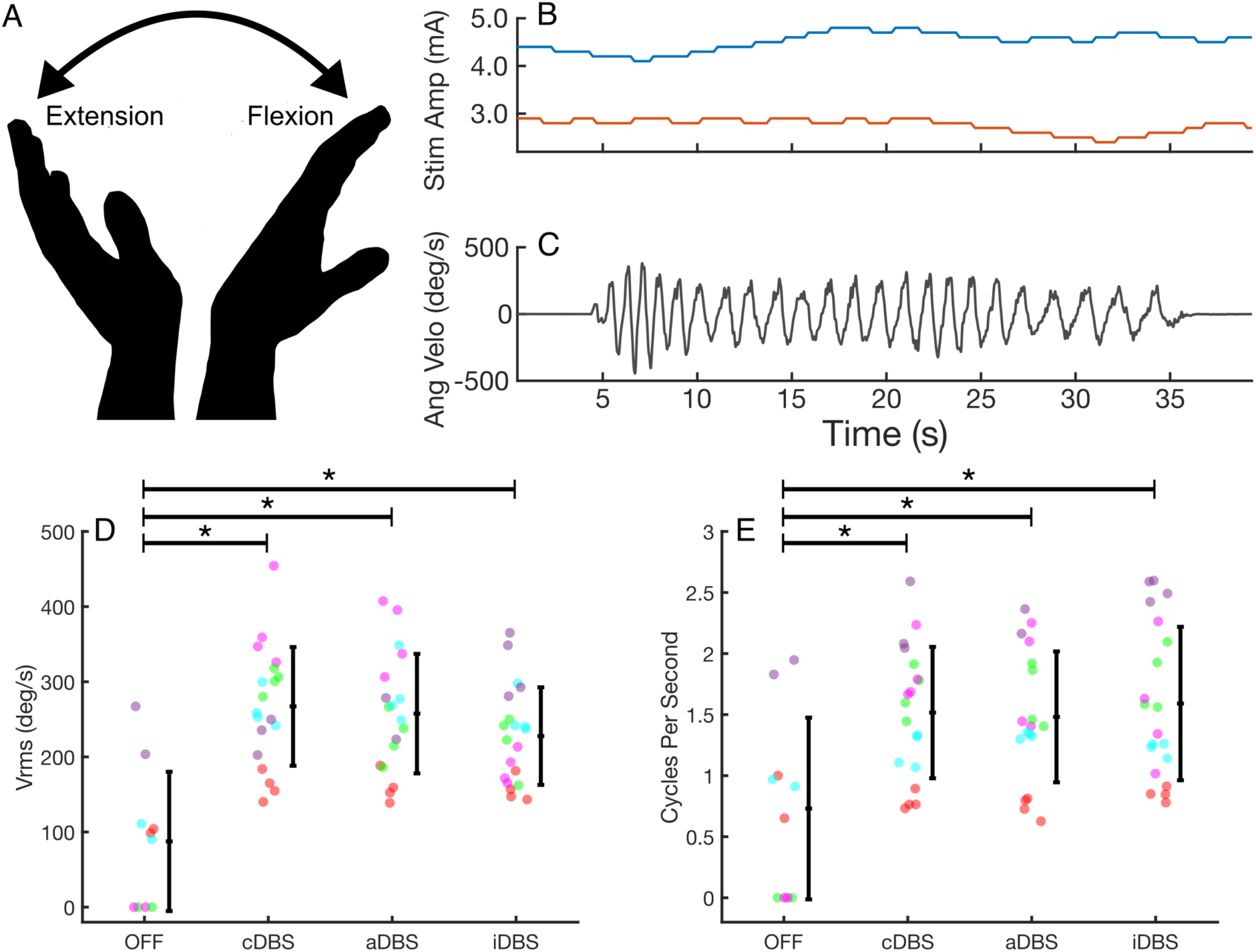
(A) Depiction of the WFE task. (B-C) Example of neural aDBS during WFE task for one participant. (B) Stimulation adapting independently for both STNs. The left STN is shown in blue and the right STN in orange. (C) Angular velocity of the hand as it flexes and extends. (D) Root mean square velocity (V_rms_) and (E) average cycles per second (Hz) during WFE across stimulation conditions. Individual data are shown, colored by participant ID, along with the average and standard deviation. * indicates p < 0.05.

There was a significant effect of stimulation condition on the number of cycles per second (F(3,70.2) = 4.96, p = 0.0035, *η*_p_^2^ = 0.18). Posthoc pairwise comparisons showed significant improvements in number of cycles per second compared to OFF stimulation for aDBS (*t* = 3.26, *p* = 0.0092), cDBS (*t* = 3.37, *p* = 0.0067), and iDBS (*t* = 3.48, *p* = 0.0047). No difference was observed between ON stimulation conditions (aDBS – cDBS: *t* = 0.014, *p* = 1.00; aDBS – iDBS: *t* = 0.11, *p* = 1.00; cDBS – iDBS: *t* = 0.10, *p* = 1.00).

### Comparison of Total Electrical Energy Delivered (TEED)

There was no significant effect of stimulation condition on TEED during the SIP task (F(2,33) = 0.0065, *p* = 0.99, 11p^2^ = 0), TBC task (F(2,52.5) = 0.067, *p* = 0.227, 11p^2^ = 0.003), or rWFE task (F(2,62) = 0.036, *p* = 0.96, 11p^2^ = 0.001).

## Discussion

The current study demonstrated the feasibility, safety, and tolerability of beta burst-driven aDBS in the STN for gait impairment and FOG in individuals with PD. A ‘bench to bedside’ model was used in which aDBS was tested first in a harnessed stepping-in-place task, followed by a FOG-provoking free walking barrier course. Neural aDBS significantly improved quantitative assessments of gait, such as percent time freezing, compared to OFF DBS and showed similar efficacy as traditional continuous DBS. Improvements in symptoms were also observed for quantitative assessments of bradykinesia and blinded clinical assessments of motor symptoms with the MDS-UPDRS III.

aDBS has been implemented in several different settings, including the peri-operative state^33–35^, in the laboratory with a chronically implanted neurostimulator^23,36^, and at home with a chronically implanted neurostimulator^37–39^. The current protocol was carried out in the laboratory setting for two primary reasons: 1. Participants included in the study suffered from gait impairment and FOG, and therefore posed a notable fall-risk. Previous aDBS studies demonstrating safety have either been at rest, during seated tasks, or in less impaired populations. Thus, it was crucial to first determine safety in a controlled lab environment; 2. No currently available neurostimulator can run embedded aDBS based on beta burst durations. The beta burst-driven aDBS used here required a ‘distributed-mode’ in which a computer-in-the-loop carried out the necessary signal processing for stimulation decisions.

The operation of aDBS has also differed across studies, with some employing rapid on/off fluctuations of stimulation in response to beta power using a single threshold^33,40^, slower adaptation in response to beta power using a dual threshold^10,23^, proportional-control^39^, or inverse algorithms based on either subcortical or cortical gamma power^37,38,41^. Here, we implemented a single-threshold algorithm where stimulation was incremented or decremented based on the duration of the most recent ongoing beta burst. However, the goal was not to necessarily trim that most recent burst, as has been done previously in the peri-operative state^33,40^; such an approach is beyond the capabilities of the current Summit RC+S neurostimulator. Instead, the most recent current burst was used as an insight into the current general state of the participant, as prolonged beta bursts are related to worse gait and motor symptoms^5,42^. Therefore, a slower ramp rate (e.g., 0.1 mA/sec) was used, rather than a rapid on/off change in stimulation. The slower ramp rate also minimized the susceptibility to artifacts in the LFP generated by changes in stimulation^22^.

Overall, aDBS significantly improved overall motor impairment compared to OFF DBS, including each sub-score, as measured by the MDS-UPDRS III. When evaluating its effect on gait specifically, aDBS significantly improved the percent time freezing and mean peak shank angular velocity (indicating faster gait speed) observed during both the harnessed SIP task and free walking TBC task. Gait asymmetry also significantly improved during SIP and showed a trend for improvement in TBC. Although gait arrhythmicity did not significantly improve, this was likely because three of the seven participants did not have asymmetry or arrhythmicity data for the OFF condition due to being frozen for the entirety of the trial. All three participants showed improvements in gait during aDBS, but this could not be captured statistically due to the lack of data for those gait metrics for the OFF condition. Meanwhile, Participant 1 (a clinical non-freezer) experienced ceiling effects for each of the gait tasks, as they demonstrated no freezing in any conditions and near normal arrhythmicity and asymmetry values. aDBS would not be expected to further improve this performance. However, bradykinesia during the WFE task and overall motor impairment, as measured by the MDS-UPDRS III) both improved substantially on aDBS for this participant (41-81% improvement in WFE; 58-70% improvement in MDS-UPDRS III), demonstrating that aDBS was effective. Interestingly, Participant 5 showed noticeably worse performance across tasks for the random iDBS control condition compared to cDBS and aDBS at both the SIP and TBC visit, suggesting that, at least for that participant, linking the adaptation to a relevant biomarker was important for improvements in behavior. iDBS was also the only ON-stimulation condition that did not significantly improve gait metrics in the harnessed SIP task, further supporting this notion. However, as a whole, iDBS did not perform significantly worse compared to cDBS or aDBS.

The aDBS control policy used here did not allow stimulation to go down to 0 mA, but instead applied an I_min_, which was the minimum level of stimulation needed to provide acceptable therapeutic benefit. The goal of this approach was to ensure that a therapeutic level of stimulation was always maintained. Previous implementations of aDBS have shown that it may not be as effective for tremor as traditional cDBS^40,43^. Whereas worse bradykinesia, rigidity, and gait are associated with increases in beta power, tremor shows the opposite phenomena where increased tremor is associated with attenuation of beta power^44^. Therefore, beta-driven aDBS has previously been vulnerable to re-emergence of tremor from the positive feedback loop between tremor, beta suppression, and drop in stimulation^23,40^. However, we saw that aDBS was notably effective for suppression of tremor across participants, likely due to the presence of the I_min_ floor. Overall, aDBS improved tremor MDS-UPDRS III sub-scores 75.4 ± 23.1%, which was even slightly higher than cDBS (70.4 ± 33.6%). The use of an I_min_, rather than allowing stimulation to go down to 0 mA, may protect against both tremor re-emergence, as well as loss of efficacy for other motor symptoms. This is supported by our finding that random iDBS, where stimulation adapted randomly within the range of I_min_ to I_max_, also showed similar efficacy as traditional cDBS. It is worth noting that the majority of previous work examining the effects of aDBS, outside of the initial Little et al., 2013 publication, have not included any sort of control condition where stimulation varies in a random fashion unlinked to the biomarker of interest. Such an inclusion is crucial for better understanding both the effects and possible mechanisms of aDBS, and we advocate for it to be included whenever possible.

A critical component of this study’s protocol was an in-depth calibration testing period to determine suitable aDBS parameters. aDBS inherently requires the setup of additional parameters compared to traditional cDBS^20,45^. We recommend the combination of quantitative assessment of behavior both OFF stimulation and ON DBS at different levels of stimulation amplitudes. Such an approach allows informed decisions of stimulation limits (i.e., I_min_ and I_max_) to ensure the maintenance of therapeutic levels of stimulation, as well as for selection of neural parameters such as the frequency band to track, value of threshold, etc. This testing allows confirmation that your signal of choice is present, modulates with stimulation, particularly in the range between I_min_ and I_max_, and is not corrupted by any sources of artifact. We carried out this testing both at rest and during movement to determine neural thresholds that were robust across movement states^46^.

### Limitations

aDBS requires a sense-friendly configuration in which the recording contacts must flank the stimulation contact/s^20^. In our cohort, four STNs across three participants required altering the clinical stimulation contacts used to ensure a sense-friendly configuration. All three of these participants were DBS re-implants, and thus further along in their disease progression and required higher stimulation amplitudes and double monopolar configurations. Typically, patients newer to DBS are more likely to only require single monopolar configurations, and thus more likely to have sense-friendly configurations. Three STNs across three participants were unusable for sensing due to either stimulation-related artifact or insufficient modulation of beta within the therapeutic window. The current cohort also demonstrated both floor and ceiling effects during the gait tasks, which may have masked some of the potential effects of aDBS. One participant was unable to perform the free walking TBC task in any condition due to the difficulty of the task and the severity of their symptoms. Similarly, several participants were frozen for the entirety of the trial in the OFF DBS condition in SIP, making it impossible to investigate any gait-related metrics such as arrhythmicity, asymmetry, etc. Meanwhile, two participants demonstrated no freezing even OFF DBS, as well as near normal gait metrics, thus leading to a ceiling effect. It is worth noting that the current study only tested the acute effects of aDBS. It is unknown from the current study whether there would be additional benefit if on the controller for longer than the two hours tested here. Lastly, the beta burst-driven aDBS control policy here required a computer-in-the-loop distributed system, and thus is limited to a research laboratory setup. Translation of such an approach would require a system capable of embedded aDBS. However, beta burst duration is correlated with beta power, and thus available systems such as the Percept PC/RC, AlphaDBS, etc., which are capable of running embedded aDBS based on power, could offer similar effects.

## Conclusion

This is the first study to demonstrate the feasibility, safety, and tolerability of beta burst-driven adaptive DBS in the STN for freezing of gait in PD. Neural aDBS improved quantitative metrics of gait compared to OFF DBS and provided similar efficacy as traditional cDBS. These improvements were maintained for all cardinal motor signs, including tremor. The current findings pave the way for future long-term testing of aDBS outside of the laboratory, even in participants with FOG and more severe motor impairments.

## Data Availability

All data produced in the present study are available upon reasonable request to the authors

## Acknowledgements

The authors would like to thank the participants who dedicated their time to this study and the Open Mind Consortium. This work was supported by NINDS UH3NS107709, U24NS113637, Robert and Ruth Halperin Foundation, John A. Blume Foundation, John E Cahill Family Foundation, and Medtronic PLC who provided the devices but no additional financial support.

## Competing Interests

The authors declare no competing interests related to this study.

## Notes

### Competing Interest Statement

The authors have declared no competing interest.

### Clinical Trial

NCT04043403

### Funding Statement

This work was supported by NINS UH3NS107709, U24NS113637, Robert and Ruth Halperin Foundation, John A. Blume Foundation, John E Cahill Family Foundation, and Medtronic PLC who provided the devices but no additional financial support.

### Author Declarations

IRB of Stanford University gave ethical approval for this work.

