## Supplementary Material for "Beta Burst-Driven Adaptive Deep Brain Stimulation Improves Gait Impairment and Freezing of Gait in Parkinson’s Disease"

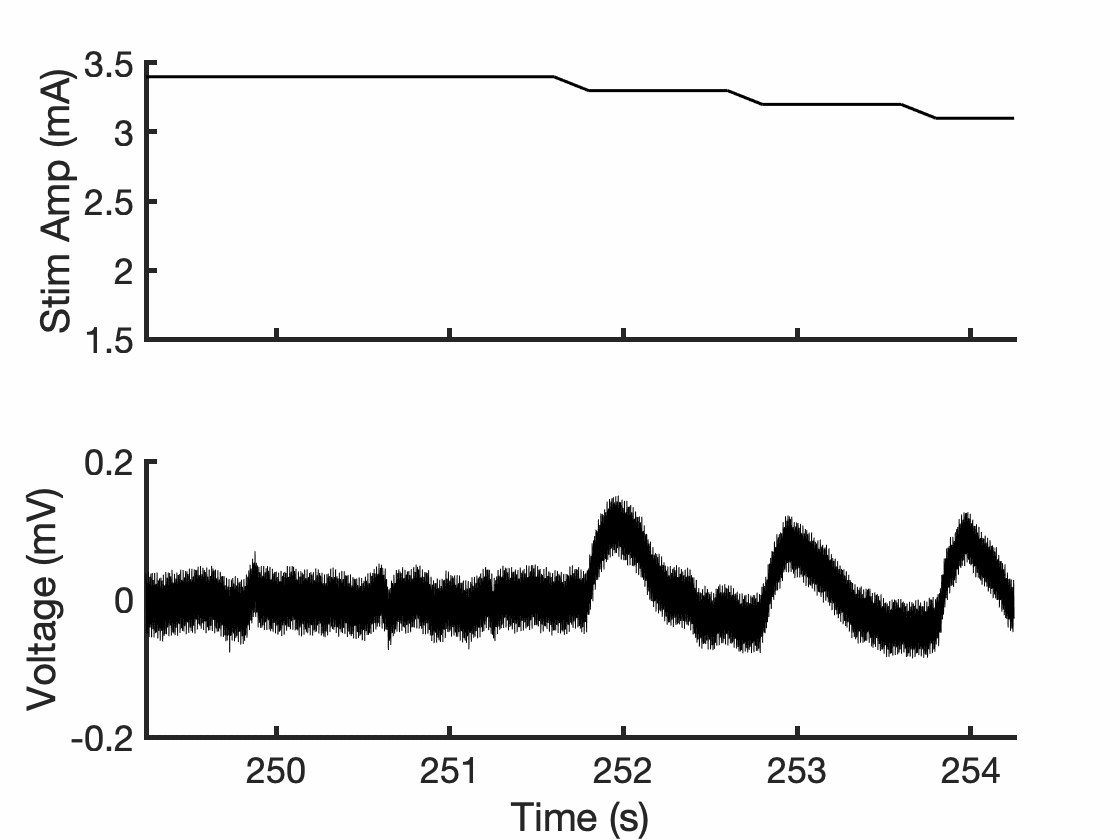

Supplementary Figure 1. Example of ramp-rate artifact in LFP. (Top) Randomized changes in stimulation amplitude during ramp rate testing. (Bottom) Corresponding LFP data. Increments/decrements of stimulation cause a transient artifact in the LFP.

Supplementary Table 1. MDS-UPDRS III ON-stimulation Sub-score Comparisons

| MDS-UPDRS III Sub-score | aDBS vs. cDBS | aDBS vs. iDBS | cDBS vs. iDBS |
| --- | --- | --- | --- |
| Bradykinesia | *t* = 0.12, *p* = 1.00 | *t* = 0.74, *p* = 0.90 | *t* = 0.58, *p* = 0.94 |
| Tremor | *t* = 0.85, *p* = 0.83 | *t* = 1.16, *p* = 0.65 | *t* = 0.31, *p* = 0.99 |
| Rigidity | *t* = 0.40, *p* = 0.98 | *t* = 1.09, *p* = 0.70 | *t* = 0.69, *p* = 0.90 |

Supplementary Table 2. Individual Gait Metrics Across Visits

| ID | Condition | SIP | | TBC (60 mins) | | TBC (120 mins) | |
| --- | --- | --- | --- | --- | --- | --- | --- |
|  |  | Arrhythmicity | Asymmetry | Arrhythmicity | Asymmetry | Arrhythmicity | Asymmetry |
| 01 | OFF | 9.1 | 17.7 | 7.5 | 7.5 | - | - |
|  | aDBS | 6.1 | 3.1 | 7.7 | 6.0 | 7.6 | 7.6 |
|  | cDBS | 5.9 | 6.9 | 6.5 | 6.5 | 7.4 | 2.7 |
|  | iDBS | 7.6 | 7.6 | 6.3 | 7.3 | 7.2 | 6.3 |
| 02 | OFF | - | - | - | - | - | - |
|  | aDBS | 13.6 | 23.8 | 7.3 | 4.2 | 7.7 | 3.0 |
|  | cDBS | 8.3 | 22.8 | 7.8 | 7.9 | 9.1 | 9.2 |
|  | iDBS | 11.8 | 24.5 | 19.3 | 22.1 | 22.4 | 35.9 |
| 03 | OFF | - | - | 24.6 | 18.3 | - | - |
|  | aDBS | 149.2 | 17.1 | 11.8 | 20.1 | 11.2 | 19.5 |
|  | cDBS | 190.4 | 11.4 | 11.6 | 12.5 | 18.1 | 7.4 |
|  | iDBS | 166.4 | 8.6 | 15.1 | 22.5 | 15.1 | 11.9 |
| 04 | OFF | 204.4 | 16.7 | 5.6 | 7.2 | - | - |
|  | aDBS | 4.7 | 3.8 | 6.9 | 7.2 | 6.5 | 10.5 |
|  | cDBS | 4.5 | 3.0 | 6.1 | 10.2 | 6.7 | 3.1 |
|  | iDBS | 4.4 | 3.0 | 5.8 | 9.2 | 5.6 | 8.3 |
| 05 | OFF | - | - | - | - | - | - |
|  | aDBS | 180.8 | 7.6 | - | - | - | - |
|  | cDBS | 131.5 | 21.6 | - | - | - | - |
|  | iDBS | - | - | - | - | - | - |
| 06 | OFF | 19.1 | 8.9 | 34.1 | 1.9 |  |  |
|  | aDBS | 16.3 | 0.4 | 41.5 | 3.7 | 28.5 | 5.5 |
|  | cDBS | 6.5 | 3.8 | 37.6 | 5.0 | 28.2 | 2.1 |
|  | iDBS | 5.6 | 3.8 | 26.0 | 4.3 | 22.8 | 9.0 |
| 07 | OFF | 10.5 | 13.7 | - | - | - | - |
|  | aDBS | 3.9 | 4.4 | - | - | - | - |
|  | cDBS | 3.6 | 6.5 | - | - | - | - |
|  | iDBS | 3.5 | 2.1 | - | - | - | - |

*Participants 05 and 07 did not complete the TBC task. Dashed lines indicate the participant was frozen for the entirety of the trial or did not complete the trial.
